# Clinical Sentiment Analysis by Large Language Models Enhances the Prediction of Hepatorenal Syndrome in Decompensated Cirrhosis

**DOI:** 10.1101/2024.11.13.24317220

**Authors:** Mason Lai, Cynthia Fenton, Jessica Rubin, Chiung-Yu Huang, Mark Pletcher, Jennifer C. Lai, Giuseppe Cullaro, Jin Ge

**Affiliations:** Department of Medicine, University of California, San Francisco; Division of Hospital Medicine, Department of Medicine, University of California, San Francisco; Division of Clinical Informatics and Digital Transformation, Department of Medicine, University of California, San Francisco; Division of Gastroenterology and Hepatology, Department of Medicine, University of California, San Francisco; Department of Epidemiology and Biostatistics, University of California, San Francisco; Division of Digestive and Liver Diseases, Department of Medicine, Columbia University

## Abstract

**Background and Aims:** Hepatorenal syndrome – Acute Kidney Injury (HRS-AKI) is a severe complication of decompensated cirrhosis that is challenging to predict. Sentiment analysis, a computational process of identifying and categorizing opinions and judgment expressed in text, may enhance traditional prediction methodologies based on structured variables. Large language models (LLMs), such as generative pretrained transformers (GPTs), have demonstrated abilities to perform sentiment analyses on non-clinical texts. We sought to determine if GPT-performed sentiment analysis could improve upon predictions using clinical covariates alone in the prediction of HRS-AKI.

**Methods:** Adult patients admitted to a single academic medical center with decompensated cirrhosis and AKI. We used a protected health information (PHI) compliant version of Microsoft Azure OpenAI GPT-4o to derive a sentiment score ranging from 0 to 1 for HRS-AKI, and conduct natural language processing (NLP) extraction of clinical terms associated with HRS-AKI in clinical notes. The area under the receiver operator curve (AUROC) was compared in logistic regression models incorporating structured variables (socio-demographics, MELD 3.0, hemodynamic parameters) with compared to without sentiment scores and NLP-extracted clinical terms.

**Results:** In our cohort of 314 participants, higher sentiment score was associated with the diagnosis of HRS-AKI (OR 1.33 per 0.1, 95% CI 1.02-1.79) in multivariate models. AUROC of the baseline model using structured clinical covariates alone was 0.639. With the addition of the GPT-4o derived sentiment score and clinical terms to structured covariates, the final model yielded an improved AUROC of 0.758 (p= 0.03).

**Conclusions:** Clinical texts contain large amounts of data that are currently difficult to extract using standard methodologies. Sentiment analysis and NLP-based variable derivation with GPT-4o in clinical application is feasible and can improve the prediction of HRS-AKI over traditional modeling methodologies alone.

## Introduction

Large language models (LLMs), such as generative pretrained transformers (GPTs) have a robust ability to understand and mimic human language^1-4^, and have been shown to be effective in a variety of tasks including predictive modeling, data extraction, and identifying diagnoses. Unstructured clinical data, such as clinical notes, contain a rich trove of information that could provide unique insights beyond structured data^5,6^. Due to the difficulty of accurately categorizing and extracting information from clinical notes at scale, traditional clinical prediction relies primarily on structured data. The rapid evolution of LLMs and their superiority in conducting natural language processing (NLP) tasks presents an unique opportunity to leverage this technology in utilizing unstructured clinical documentation^7^.

In particular, sentiment analysis is an NLP technique in which free text is computationally analyzed to derive a numeric score that conveys the polarity (positive or negative) attitude, consensus, or feeling for a given sample of unstructured text data. Sentiment analysis has historically been done using lexicon-based techniques (which rely on predefined dictionaries of words with positive or negative annotated values) or machine learning methods such as support vector models or neural networks^8,9^. Prior research has explored using medical sentiment to predict clinical outcomes, such as short-term survival in hospitalized patients^10-12^. In contrast to traditional sentiment analysis methods, LLMs, which are pretrained on large corpuses of text, have demonstrated versatile NLP abilities, such as information extraction and categorization. Moreover, they have also been widely deployed in the technology industry, such as in social media or e-commerce applications, to conduct sentiment analyses on user comments and reviews to give an overall summary or consensus^13,14^. When applied to clinical text, sentiment analysis, may be able to better determine the aggregate opinions and consensus of clinicians and enhance clinical prediction modeling.

Hepatorenal syndrome-acute kidney injury (HRS-AKI) is the one of the deadliest complications of decompensated cirrhosis, approaching a 50% mortality rate at 30-90 days^15,16^. While clinically important, predictors of HRS-AKI have not been reliably identified in the literature – although some covariates with more consistent signal include ascites, lab markers of cirrhosis, and spontaneous bacterial peritonitis^17,18^. The reason why traditional prediction models have failed to be effective for HRS-AKI is because its diagnosis is heavily based on clinical judgement. Sentiment analysis, as implemented through LLMs and applied to clinical documentation, therefore, may offer unique value in improving predictions.

In the present study, we sought to leverage these capabilities and utilize our institution’s protected health information (PHI) compliant version of GPT-4o to conduct sentiment analysis on clinical documentation and determine whether this approach improves model prediction of HRS-AKI over traditional clinical covariates alone.

## Methods

### Study Design and Population

This is a retrospective sub-analysis of the prospective Functional Assessment in Liver Transplantation (FrAILT) study who were consecutively hospitalized at the University of California, San Francisco (UCSF) from August 2012 to September 2023. In brief, the FrAILT study consists of a cohort of adult (age 18+) patients with decompensated cirrhosis who were listed for liver transplantation. Specifically, this subset consists of 314 patients with decompensated cirrhosis who were admitted to UCSF Medical Center with acute kidney injury (AKI) at the time of admission, defined as an increase in serum creatinine of ≥0.3mg/dL or ≥50% from their last known or presumed baseline in the prior 7 days. Patients on dialysis as an outpatient, or without an accessible history and physical (H&P) admission note were excluded from the study. This study was approved by the UCSF IRB under IRB # 11-07513. Funding sources had no role in the study design, data collection, or manuscript.

### Primary Predictor: Sentiment Analyses and Score

Sentiments within admission history and physicals from the primary team were extracted using the “Versa” API, which is the PHI and intellectual property compliant deployment of Microsoft Azure OpenAI GPT family of LLMs deployed at UCSF. We utilized gpt-4o-2024-08-06, version dated as of August 6, 2024 for all analyses. We conducted iterative prompt engineering on a subset of clinical notes until the desired output was achieved^19^. This desired output was defined as gpt-4o producing the following outputs:

1. Numeric score between 0-1 for clinician sentiment for HRS-AKI with an explanation of the logic,
2. Five non-overlapping key phrases or terms used in the sentiment score generation,
3. Binary variable indicating the presence or absence of specific NLP-extracted terms in the text.

The final engineered system prompt stated that the LLM is an expert in both liver and kidney disease with expertise in identifying hepatorenal syndrome (HRS-AKI), and with a background in sentiment analysis (Supplemental Materials C). The final engineered user prompt requested that the GPT read the provided note and generate a sentiment value between 0 and 1, based on whether the clinical team felt the patient had HRS-AKI. Scores closer to 1 indicated higher suspicion for HRS-AKI.

### Secondary Predictor: NLP-Extracted Clinical Terms

NLP is a technique that involves extracting meaning or values from unstructured text, which can then be used in other machine-learning methods. Given the ability of generative AI to understand and mimic human language, we sought to use gpt-4o to extract aspects of free-text that are challenging to quantify in structured data. Specifically, we prompted gpt-4o to identify if the patient had 1) history of recurrent AKI from diuretics; 2) signs concerning for shock including vasopressor use; 3) and refractory ascites. Prompts for free-text extraction are shown in Supplemental Materials D. These variables, here forth referred to as “NLP-extracted clinical terms” were added as covariates to logistic regression and assessed for association with HRS-AKI diagnosis.

### Structured Data Covariates

The structured covariate set used in the initial clinical model was defined *a priori*. These included sociodemographic variables (age, sex), mean arterial pressure, and serum lab values at the time of admission (albumin, total bilirubin, international normalized ratio [INR], sodium), serum creatinine at the time of admission, baseline serum creatinine, and presence of ascites. These data were derived from the UCSF electronic health record (EHR) clinical data warehouse. All considered covariates were included in the model, irrespective of their p-value.

### Primary Outcome

We identified patients who developed hepatorenal syndrome based on the discharge summaries for each encounter. We defined HRS-AKI according to current International Club of Ascites and Acute Disease Quality Initiative guidelines^20^, specifically:

a. Cirrhosis with ascites,
b. Increase in serum creatinine ≥0.3 mg/dl (26.5 μmol/L) within 48 h or ≥50% from baseline value, known or presumed, to have occurred within the prior 7 days and/or urine output ≤0.5 ml/kg for6h,
c. Absence of improvement in serum creatinine and/or urine output within 24 h following adequate volume resuscitation (when clinically indicated), and
d. Absence of strong evidence for an alternative explanation as the primary cause of AKI.

Given that hepatorenal syndrome is a challenging diagnosis to make, we fed the discharge summaries to gpt-4o, and prompted it to identify if the patient was diagnosed with hepatorenal syndrome. We completed manual validation on a subset of patients. The sample size (n=231) was determined with a 95% confidence level, a target of 90% sensitivity, and a 10% margin of error in sensitivity and specificity, and a 15% prevalence of HRS-AKI^21^. These prompts are reported in Supplemental Materials E. Test characteristics are reported in Table 4.

### Statistical Analysis

We chose to utilize logistic regression given its ease of interpretability. Univariate analyses were first conducted with each covariate for association with HRS-AKI. We generated four different multivariate models:

1. The first included structured clinical data only, taken from the day of admission, including sociodemographic variables, hemodynamics, lab values, and presence or absence of ascites.
2. The second model included all structured data and added the sentiment value for HRS-AKI generated by gpt-4o.
3. The third model included all structured data, and indicator variables diuretic intolerance, refractory ascites, and shock that were derived from NLP-extracted clinical terms.
4. The fourth model included all structured data, the sentiment value for HRS-AKI, and indicator variables for diuretic intolerance, refractory ascites, and shock that were derived from free-text using NLP.

We split the dataset into 80%/20% training/testing (n=253 and 56, respectively). Statistical analyses were completed in R 4.2.2. The ‘pROC’ v1.18.5 package was heavily utilized in our analyses. All statistical testing used a two-sided p-value cutoff of 0.05. With respect to test performance characteristics, specificity, sensitivity, positive predictive value, negative predictive value, and area under the receiver operator curves (AUROC) were used to evaluate model performance. Cutoff values were determined using Youden’s index. AUROC curves were compared using DeLong’s test, a nonparametric test for differences in ROCs^22^. All analyses and results reported below used the temperature of 0.0 (a parameter which controls the randomness of the responses) in gpt-4o as this is the most deterministic setting in gpt-4.

## Results

### Study population

Of the 314 participants, median age was 59.5 (Q1-Q3: 52.3-64.1) years and was similar between those who experienced HRS-AKI and those with non-HRS AKI (Table 1). Fifty two percent of the overall cohort was male. Median baseline creatinine was 1.44 (Q1-Q3: 1.05-1.93) in those with HRS-AKI and 1.09 (Q1-Q3: 0.84-1.47) in those with non-HRS AKI. Median MELD 3.0 was 34 (Q1-Q3: 33-37) in those with HRS-AKI and 31 (Q1-Q3: 27-35) in those with non-HRS-AKI. Median sentiment score was 0.8 (Q1-Q3: 0.7-0.9) in those who experienced HRS-AKI compared to 0.7 (Q1-Q3: 0.6-0.8) in those who did not experience HRS-AKI.

**Table 1:**
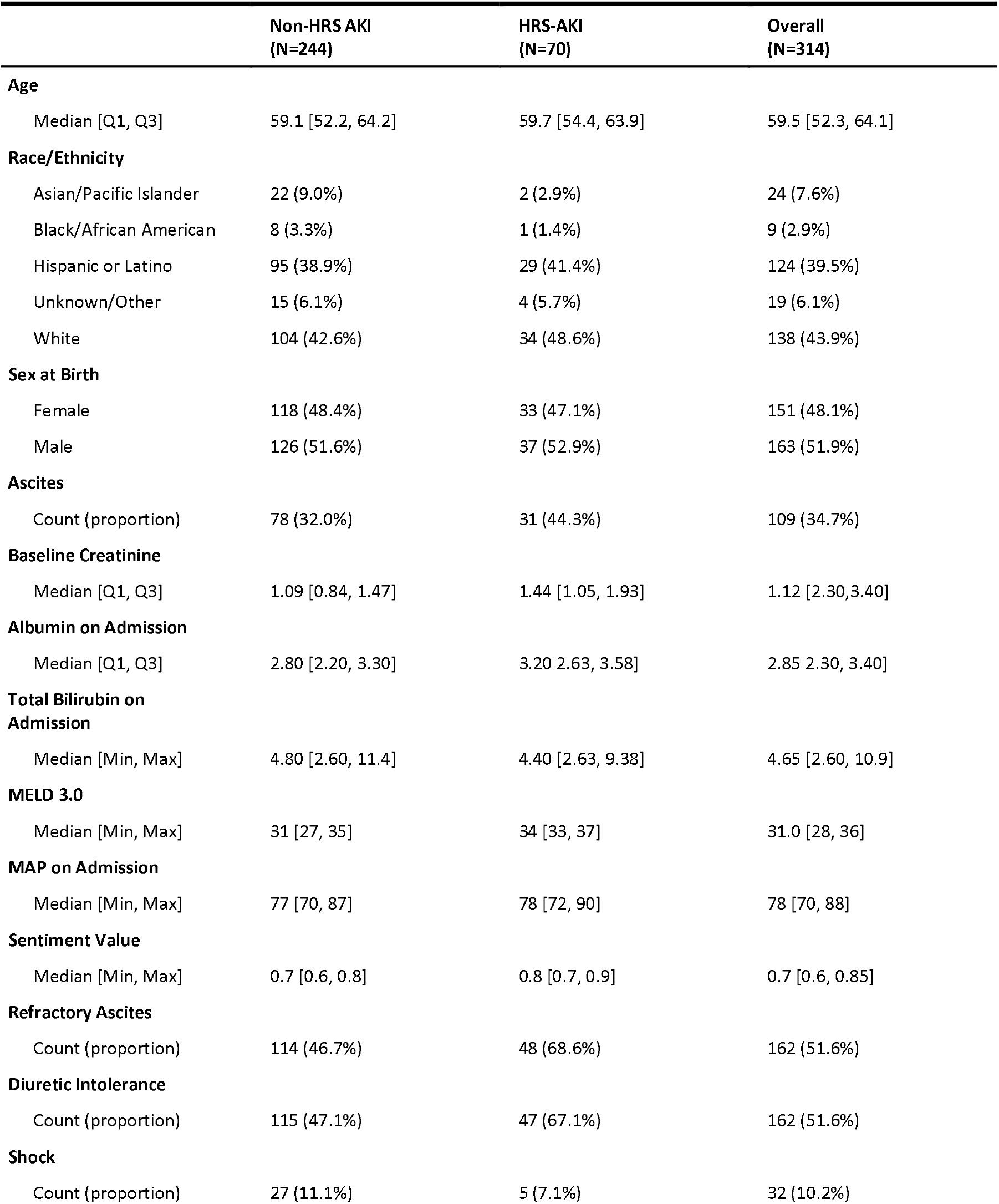
Participant characteristics by HRS-AKI status.

### Multivariate Model 1: Association Between Structured Clinical Covariates and HRS-AKI

The clinical covariates in Table 2 were evaluated in univariate logistic regression for association with HRS-AKI within the training data. Serum creatinine at baseline (OR 1.61, 95%CI 1.19-2.26), at the time of diagnosis of AKI (OR 1.73, 95%CI 1.38-2.23), serum albumin (OR 1.65, 95%CI 1.15-2.38), and total bilirubin (OR 0.95, 95%CI 0.91-0.99)) were significantly associated with HRS-AKI, while all other clinical covariates were not significant. In multivariate regression adjusted for other clinical covariates, serum creatinine at AKI on admission (OR 1.81, 95%CI 1.22-2.69) and presence of ascites (OR 2.02, 95%CI 1.06-3.87) were associated with HRS-AKI.

**Table 2:** Associations of sentiment scores on hepatorenal syndrome in patients with decompensated cirrhosis hospitalized with AKI.

|  | Univariate |  | Multivariate Model 1† |  | Multivariate Model 2†† |  | Multivariate Model 3‡ |  | Multivariate Model 4§ |  |
| --- | --- | --- | --- | --- | --- | --- | --- | --- | --- | --- |
|  | OR (95% CI) | p-value | OR (95% CI) | p-value | OR (95% CI) | p-value | OR (95% CI) | p-value | OR (95% CI) | p-value |
| Sentiment score (per 0.1 increase) | 1.58 (1.27-2.04) | <0.001 | - | - | 1.37 (1.08-1.80) | 0.01 | - | - | 1.33 (1.02-1.79) | 0.05 |
| Shock | 0.67 (0.22-1.70) | 0.43 | - | - | - | - | 0.45 (0.11-1.51) | 0.23 | 0.35 (0.09-1.21) | 0.35 |
| Refractory Ascites | 2.66 (1.45-5.03) | <0.01 | - | - | - | - | 2.34 (1.08-5.23) | 0.03 | 1.86 (0.83-4.30) | 0.14 |
| Diuretic Intolerance | 2.26 (1.26-4.17) | 0.01 | - | - | - | - | 1.58 (0.78-3.29) | 0.21 | 1.35 (0.64-2.86) | 0.44 |
| **Clinical Covariates:** |  |  |  |  |  |  |  |  |  |  |
| Male Sex | 0.92 (0.52, 1.63) | 0.77 | 0.79 (0.40, 1.52) | 0.48 | 0.72 (0.36-1.41) | 0.34 | 0.88 (0.44, 1.76) | 0.72 | 0.83 (0.41, 1.68) | 0.60 |
| Age (per 1 year) | 1.02 (0.99, 1.06) | 0.14 | 1.00 (0.96, 1.04) | 0.96 | 1.00 (0.96-1.04) | 0.97 | 1.00 (0.96, 1.04) | 0.94 | 1.00 (0.96, 1.04) | 0.94 |
| Albumin (per 1 mg/dL) | 1.65 (1.15, 2.38) | 0.01 | 1.36 (0.90, 2.05) | 0.14 | 1.21 (0.79, 1.85) | 0.38 | 1.17 (0.75, 1.83) | 0.48 | 1.11 (0.71, 1.76) | 0.64 |
| Sodium (per 1 mg/dL) | 0.99 (0.95, 1.04) | 0.73 | 0.96 (0.91, 1.01) | 0.15 | 0.96 (0.91, 1.02) | 0.20 | 0.97 (0.92, 1.03) | 0.33 | 0.97 (0.91, 1.03) | 0.26 |
| INR (per 1) | 1.01 (0.67, 1.47) | 0.96 | 1.13 (0.70, 1.78) | 0.59 | 1.00 (0.61, 1.60) | 0.99 | 1.26 (0.74, 2.08) | 0.38 | 1.21 (0.70, 2.05) | 0.47 |
| Total Bilirubin (per 1 mg/dL) | 0.95 (0.91, 0.99) | 0.03 | 0.97 (0.91, 1.02) | 0.24 | 0.96 (0.91, 1.01) | 0.18 | 0.98 (0.92, 1.03) | 0.43 | 0.96 (0.90, 1.01) | 0.20 |
| MAP (per 1 mm/Hg) | 1.01 (0.99, 1.03) | 0.34 | 1.01 (0.99, 1.04) | 0.31 | 1.01 (0.99, 1.04) | 0.30 | 1.01 (0.99, 1.04) | 0.25 | 1.01 (0.99, 1.04) | 0.29 |
| Baseline Cr (per 1 mg/dL) | 1.61 (1.19, 2.26) | <0.01 | 0.82 (0.48, 1.45) | 0.49 | 0.81 (0.47, 1.44) | 0.81 | 0.62 (0.34, 1.14) | 0.12 | 0.62 (0.33, 1.15) | 0.12 |
| Cr at AKI (per 1 mg/dL) | 1.73 (1.38, 2.23) | <0.01 | 1.81 (1.22, 2.69) | <0.01 | 1.69 (1.13, 2.53) | 0.01 | 2.18 (1.43, 3.39) | <0.01 | 1.99 (1.30, 3.10) | <0.01 |
| Ascites Present | 1.77 (0.98, 3.18) | 0.06 | 2.02 (1.06, 3.87) | 0.03 | 1.95 (1.01, 3.77) | 0.05 | 1.97 (1.00, 3.89) | 0.05 | 2.09 (1.05, 4.18) | 0.03 |

† Includes age, sex, serum albumin, serum sodium, INR, total bilirubin, mean arterial pressure, baseline creatinine, presence of ascites.
†† Includes age, sex, serum albumin, serum sodium, INR, total bilirubin, mean arterial pressure, baseline creatinine, presence of ascites, sentiment score.
‡ Includes age, sex, serum albumin, serum sodium, INR, total bilirubin, mean arterial pressure, baseline creatinine, presence of ascites, NLP derived variables: shock, refractory ascites, diuretic intolerance.
§ Includes age, sex, serum albumin, serum sodium, INR, total bilirubin, mean arterial pressure, baseline creatinine, presence of ascites, sentiment score, NLP derived variables: shock, refractory ascites, diuretic intolerance.

### Multivariate Model 2: Association Between Structured Clinical Covariates + Sentiment Score and HRS-AKI

We ran logistic regressions within the training data using the sentiment scores generated by LLM and assessed the association with HRS-AKI diagnosis. Sentiment scores were predefined as a score from 0 to1. In univariate analysis, higher sentiment score was associated with higher odds (OR 1.58 per 0.1 higher score, 95%CI 1.27-2.04) of incident HRS-AKI (Table 2). After adjustment for clinical covariates including sociodemographic factors, MELD score components, serum creatinine, and hemodynamic parameters, higher sentiment score retained association with HRS-AKI (OR 1.37 per 0.1 higher score, 95%CI 1.08-1.80). After additional adjustment for NLP derived variables diuretic intolerance, refractory ascites, and shock, higher sentiment score remained associated with HRS-AKI (OR 1.33 per 0.1 higher score, 95%CI 1.02-1.79).

To gain insight into how gpt-4o was generating sentiment scores, we prompted the LLM to identify five words or phrases that it thought was most influential in assigning its sentiment score. The top 10 most frequently cited words are listed in Table 3. Most of these words reflect an aspect of AKI or severity of cirrhosis. “Albumin”, “Ascites”, and “Renal” were also among the most frequently cited words, indicating the model may have been using some typical HRS-AKI risk factors and treatment to guide its decision when assigning a sentiment score. Notably, alternative AKI etiologies were not among the most mentioned words. Specifically, there was no mention of acute glomerular injury, acute tubular necrosis, granular casts, hematuria, obstruction, nephrotoxins, or shock, which would point to an alternative etiology for AKI.

**Table 3:** High frequency words used by LLM to generate sentiment score.

|  | Count (%) |
| --- | --- |
| Albumin | 148 (42.7%) |
| Ascites | 103 (29.7%) |
| HRS | 81 (23.3%) |
| Diuretics | 69 (19.9%) |
| Refractory | 56 (16.1%) |
| Renal | 49 (14.1%) |
| Midodrine | 47 (13.5%) |
| AKI | 46 (13.3%) |
| Cirrhosis | 46 (13.3%) |
| Creatinine | 41 (11.8%) |
| Challenge | 37 (10.7%) |

**Table 4:**
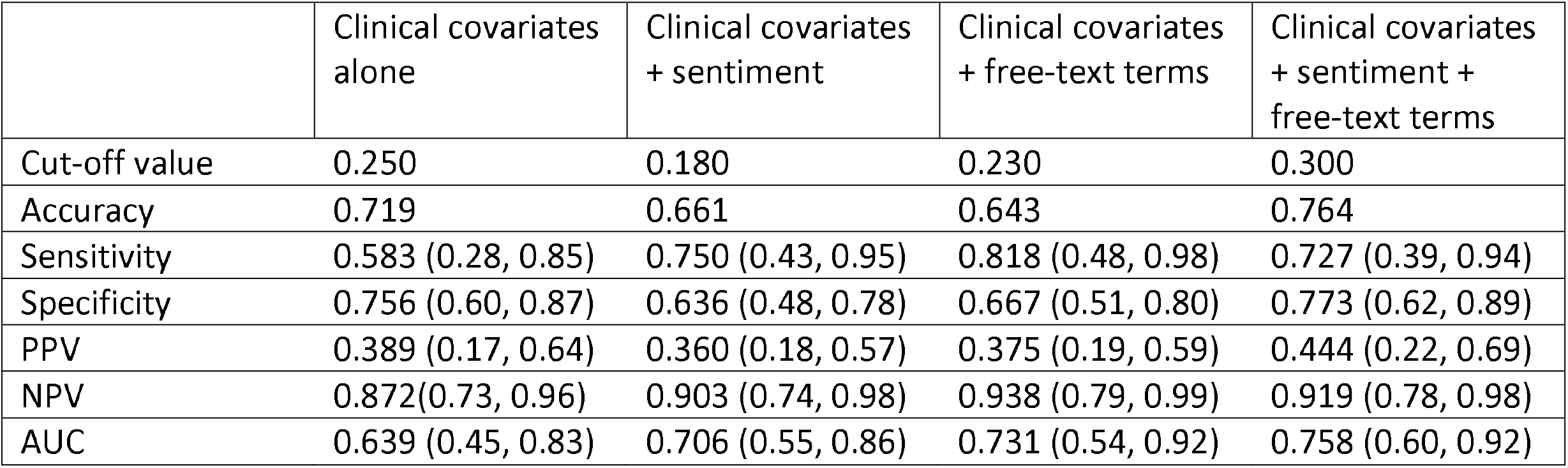
Predictive performance of models for HRS-AKI.

### Multivariate Model 3: Association Between Structured Clinical Covariates + NLP-Extracted Clinical Terms and HRS-AKI

In univariate analysis, refractory ascites as compared to lack of refractory ascites was significantly associated with HRS-AKI (OR 2.66, 95%CI 1.45-5.03), as was history of diuretic intolerance (OR 2.26, 95%CI 1.26-4.17). After adjustment for clinical covariates, refractory ascites and diuretic intolerance were both attenuated and no longer significant although maintained the direction of association (OR 1.86, 95%CI 0.83-4.30, and OR 1.35, 95%CI 0.64-2.86, respectively). Shock was not significantly associated with HRS-AKI in univariate or multivariate models.

### Multivariate Model 4: Associations Between Structured Clinical Covariates + Sentiment Scores + NLP-Extracted Clinical Terms and HRS-AKI

In the final multivariate model, sentiment scores, NLP-extracted clinical terms, and structured clinical covariates were included. In this model, sentiment scores remained associated with the development of HRS-AKI (OR 1.33, 95%CI 1.02-1.79), while the NLP-extracted clinical terms for shock, refractory ascites, and diuretic intolerance were attenuated. Of the clinical covariates, serum creatinine at the time of AKI (OR 1.99, 95%CI 1.30-3.10) and presence of ascites (OR 2.09, 95%CI 1.05-4.18) remained significantly associated with HRS-AKI.

### Test Characteristics

Sensitivity, specificity, positive predictive value, negative predictive value, accuracy, and AUROC (Figure 1) were used to compare performance of the regression models (Table 4). Sequential improvements were seen in the AUROC of the models with additions from the free-text admission history and physical. The AUROC for the initial multivariate clinical model was 0.638. Addition of sentiment score to this model yielded an AUROC of 0.706 (p-value 0.09 compared to clinical model by Delong’s test). NLP terms added to the model yielded an AUROC of 0.731 (p-value 0.14). The AUROC for the final model including traditional clinical covariates, sentiment score, and free-text derived factors was 0.758 (p-value 0.03 compared to clinical model by DeLong’s test).

**Figure 1:**
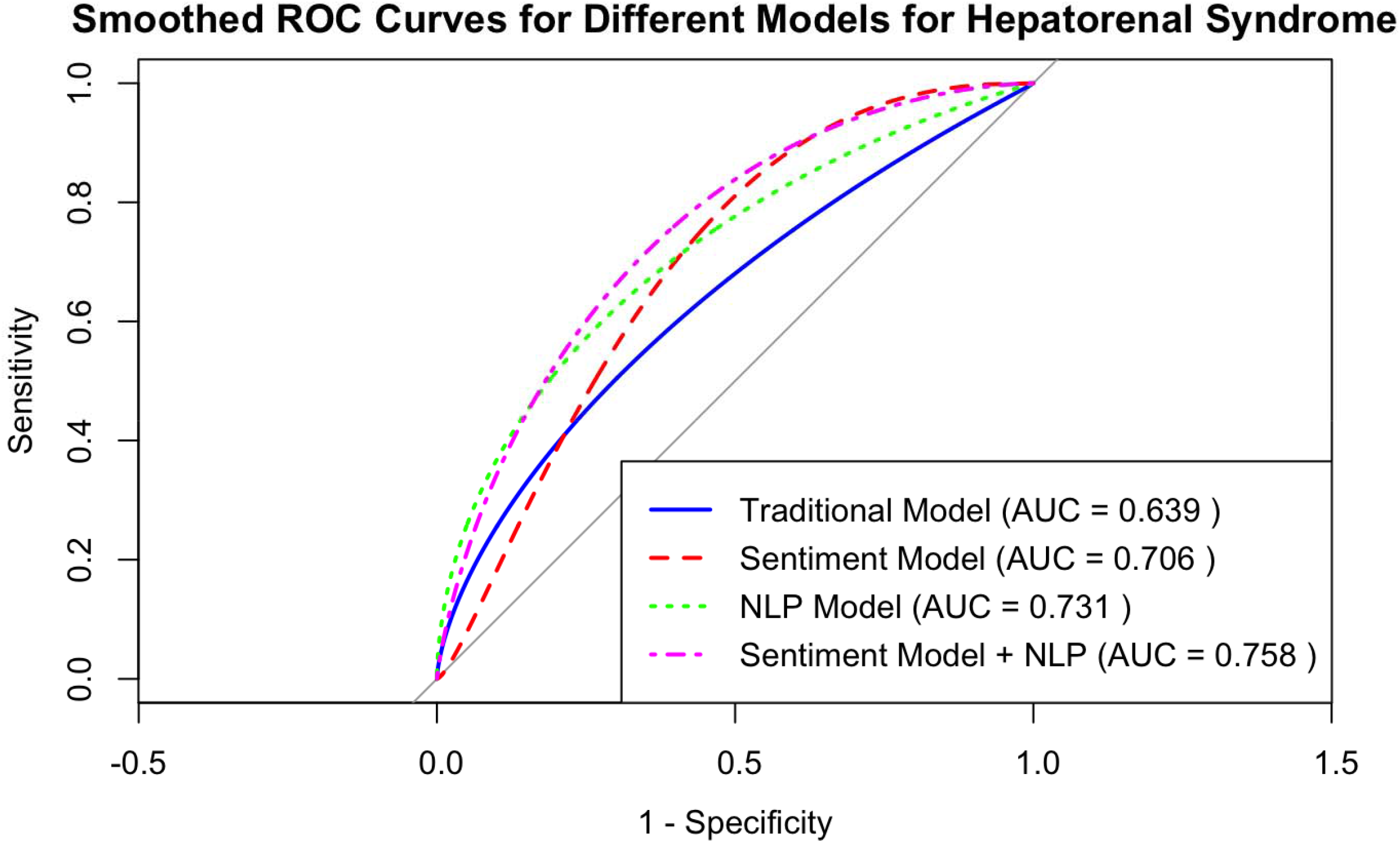

## Discussion

In this present study, we utilized LLMs, specifically gpt-4o, to perform clinical sentiment analyses and extract variables from unstructured notes to enhance the prediction of a clinically challenging diagnosis – HRS-AKI. To our knowledge, this study is the first of its kind to utilize LLM to perform clinical sentiment analyses in hepatology. Notably, these AI-derived inputs were added to traditional clinical covariates to determine if they improved model prediction for hepatorenal syndrome using logistic regression. We found that sentiment score and NLP derived variables for diuretic intolerance, refractory ascites and shock improved model prediction, compared to traditional covariates alone (AUROC 0.758 compared to 0.639). Given the historical difficulty in utilizing unstructured text for clinical research, these techniques enabled by generative AI represent a new paradigm for clinical predictive modeling. Clinicians spend a substantial amount of time and cognitive effort generating unstructured data, much of it in the form of assessments and plans in hospital admission notes. This data represents a trove of information, providing unique insights into patient status and risks that traditional structured variables overlook^5^. This data has historically been largely incompatible with traditional machine learning methods for predictive model development, given its unstructured nature. With the recent developments in generative AI^1^, it is naturally positioned to interpret and summarize clinician sentiment for use in prediction model building.

Sentiment scores have been used to leverage clinician sentiment in prior research using conventional NLP techniques and has been shown to predict hard outcomes including mortality^11,12^. In the case of HRS-AKI, early diagnosis is both challenging and imperative given the high mortality associated with the condition. The sentiment scores derived by generative AI technologies in the present study likely reflect a combination of clinician concern and therapeutic intent, enabling it to capture a unique aspect of HRS-AKI likelihood. Indeed, when asked to present its logic for the derivation of sentiment scores in the form of highly influential phrases in the admission note, the three most frequent words cited were 1) “albumin” - likely reflecting albumin challenge, 2) “ascites” – reflecting one of the criteria for HRS-AKI diagnosis, and 3) “HRS” – reflecting differential diagnosis and clinical suspicion in the assessment of AKI. Despite this, the nature of derivation of these sentiment scores remains relatively opaque, and it is unclear if these terms are the main drivers in deriving the sentiment score, or if the LLM is simply reproducing phrases related to HRS-AKI.

To build upon and further explore the NLP potential of LLMs, we also used gpt-4o to derive variables that are embedded within the unstructured text, which has previously been shown in multiple studies to have excellent concordance with manual chart review^2^. Specifically, we used the admission note and focused on variables related to ascites and shock, both of which are challenging to categorize in structured data. For example, a history of diuretic intolerance due to recurrent AKI is not an International Classification of Diseases, Revision 10 diagnosis code, and thus challenging to reliably code into structured data. We hypothesized that individuals with recurrent AKI due to diuretics would be highly sensitive to hemodynamic changes and thus at greater risk for HRS-AKI.

Future studies might try to leverage generative AI to derive sentiment scores to identify groups of patients that are intractable for identification to traditional methodologies. As an example, one might use sentiment analysis on social work and clinician notes to identify patients who are at higher risk of being lost to follow-up, allowing targeted interventions to improve follow-up rates. As a research application, while numerous validated scoring systems exist for sepsis, it remains largely a clinical diagnosis – an AI-derived sentiment score may enable capture of clinician suspicion for sepsis beyond ICD-10 codes and scoring systems, and thus improve identification of the condition for use in research. Notably, we used an off-the-shelf configuration of ChatGPT 4o. Future applications might assess if using retrieval augmented generation might improve the sensitivity and specificity of derived sentiment scores and free-text derived indicator variables, and by extension increase its impact on prediction augmentation.

The present study has several limitations. First, the study was conducted on a relatively small cohort at a single tertiary liver transplant center. External validation is needed to determine if the results are generalizable to other contexts. Second, sentiment scores and NLP-derived indicators were drawn from admission history and physical notes, in which there is significant variability in the level of detail. Future research should examine if the sentiment score and/or NLP-derived indicators are dependent on either quality and/or quantity of notes. Third, the outcome was principally derived with gpt-4o review of discharge summaries, while this has excellent concordance with manual chart review, there is potential for measurement bias. Fourth, with use of LLMs, there is a risk of hallucination in the output^3^, however, this phenomenon is likely random and likely to result in a bias towards the null.

In conclusion, generative AI can conduct sentiment analyses and derive sentiment scores from unstructured clinical notes. These sentiment scores captured nuanced information that exceeded the insights available from structured data alone, and have the potential to improve prediction models. Furthermore, generative AI can derive difficult to quantify risk factors from unstructured data, such as recurrent AKI from diuretics. These NLP derived parameters work synergistically with traditional structured data when applied to outcome prediction such as HRS-AKI in patients with decompensated cirrhosis.

## Data Availability

Data is available upon reasonable request at the discretion and approval of the authors.

## Author Contributions

Authorship was determined using ICMJE recommendations.

**Lai, M:** Concept and design; data extraction, analysis and interpretation of data, drafting of manuscript, critical revision of the manuscript for important intellectual content

**Fenton:** Data extraction, critical revision of the manuscript for important intellectual content Rubin: Critical revision of the manuscript for important intellectual content

**Huang:** Critical revision of the manuscript for important intellectual content

**Pletcher:** Critical revision of the manuscript for important intellectual content, study supervision

**Lai, J:** Critical revision of the manuscript for important intellectual content, study supervision

**Cullaro:** Concept and design, analysis and interpretation of data, drafting of manuscript, critical revision of the manuscript for important intellectual content, study supervision

**Ge:** Concept and design, analysis and interpretation of data, drafting of manuscript, critical revision of the manuscript for important intellectual content, study supervision

## Writing Assistance

None

## Acknowledgements

The authors thank the UCSF AI Tiger Team, Academic Research Services, Research Information Technology, and the Chancellor’s Task Force for Generative AI for their software development, analytical, and technical support related to the use of Versa API gateway (the UCSF secure implementation of large language models and generative AI by means of API gateway), Versa chat (the chat user interface), and related data assets.

## References

1. Dave T, Athaluri SA, Singh S. ChatGPT in medicine: an overview of its applications, advantages, limitations, future prospects, and ethical considerations. Mini Review. Frontiers in Artificial Intelligence. 2023-May-04 2023;6 doi:10.3389/frai.2023.1169595

2. Ge J, Li M, Delk MB, Lai JC. A comparison of large language model versus manual chart review for extraction of data elements from the electronic health record. medRxiv. Sep 4 2023; doi:10.1101/2023.08.31.23294924

3. Nayak A, Alkaitis MS, Nayak K, Nikolov M, Weinfurt KP, Schulman K. Comparison of History of Present Illness Summaries Generated by a Chatbot and Senior Internal Medicine Residents. JAMA Internal Medicine. 2023;183(9):1026–1027. doi:10.1001/jamainternmed.2023.2561

4. Han C, Kim DW, Kim S, et al. Evaluation of GPT-4 for 10-year cardiovascular risk prediction: Insights from the UK Biobank and KoGES data. iScience. Feb 16 2024;27(2):109022. doi:10.1016/j.isci.2024.109022

5. Velupillai S, Suominen H, Liakata M, et al. Using clinical Natural Language Processing for health outcomes research: Overview and actionable suggestions for future advances. J Biomed Inform. Dec 2018;88:11–19. doi:10.1016/j.jbi.2018.10.005

6. Demner-Fushman D, Elhadad N. Aspiring to Unintended Consequences of Natural Language Processing: A Review of Recent Developments in Clinical and Consumer-Generated Text Processing. Yearbook of Medical Informatics. 2016;25:224–233.

7. Ge J, Lai JC. Artificial intelligence-based text generators in hepatology: ChatGPT is just the beginning. Hepatol Commun. Apr 1 2023;7(4) doi:10.1097/hc9.0000000000000097

8. Nandwani P, Verma R. A review on sentiment analysis and emotion detection from text. Soc Netw Anal Min. 2021;11(1):81. doi:10.1007/s13278-021-00776-6

9. Denecke K, Reichenpfader D. Sentiment analysis of clinical narratives: A scoping review. Journal of Biomedical Informatics. 2023/04/01/ 2023;140:104336. doi:10.1016/j.jbi.2023.104336

10. Denecke K, Deng Y. Sentiment analysis in medical settings: New opportunities and challenges. Artificial Intelligence in Medicine. 2015/05/01/ 2015;64(1):17–27. doi:10.1016/j.artmed.2015.03.006

11. Gao Q, Wang D, Sun P, Luan X, Wang W. Sentiment Analysis Based on the Nursing Notes on In-Hospital 28-Day Mortality of Sepsis Patients Utilizing the MIMIC-III Database. Comput Math Methods Med. 2021;2021:3440778. doi:10.1155/2021/3440778

12. Waudby-Smith IER, Tran N, Dubin JA, Lee J. Sentiment in nursing notes as an indicator of out-of-hospital mortality in intensive care patients. PLoS One. 2018;13(6):e0198687. doi:10.1371/journal.pone.0198687

13. Seong B, Song K. Sentiment analysis of online responses in the performing arts with large language models. Heliyon. Dec 2023;9(12):e22457. doi:10.1016/j.heliyon.2023.e22457

14. Chu M, Chen Y, Yang L, Wang J. Language interpretation in travel guidance platform: Text mining and sentiment analysis of TripAdvisor reviews. Front Psychol. 2022;13:1029945. doi:10.3389/fpsyg.2022.1029945

15. Patidar KR, Belcher JM, Regner KR, et al. Incidence and outcomes of acute kidney injury including hepatorenal syndrome in hospitalized patients with cirrhosis in the US. Journal of Hepatology. 2023;79(6):1408–1417. doi:10.1016/j.jhep.2023.07.010

16. Alessandria C, Ozdogan OC, Guevara Mn, et al. LIVER FAILURE AND LIVER DISEASE. 2005:

17. Ginès A, Escorsell A, Ginès P, et al. Incidence, predictive factors, and prognosis of the hepatorenal syndrome in cirrhosis with ascites. Gastroenterology. Jul 1993;105(1):229–36. doi:10.1016/0016-5085(93)90031-7

18. Janíčko M, Veseliny E, Senajová G, Jarčuška P. Predictors of hepatorenal syndrome in alcoholic liver cirrhosis. Biomedical papers of the Medical Faculty of the University Palacky, Olomouc, Czechoslovakia. 2015;159 4:661–5.

19. Ge J, Chen IY, Pletcher MJ, Lai JC. Prompt Engineering for Generative Artificial Intelligence in Gastroenterology and Hepatology. Official journal of the American College of Gastroenterology | ACG. 2024;119(9)

20. Nadim MK, Kellum JA, Forni L, et al. Acute kidney injury in patients with cirrhosis: Acute Disease Quality Initiative (ADQI) and International Club of Ascites (ICA) joint multidisciplinary consensus meeting. Journal of Hepatology. 2024;81(1):163–183. doi:10.1016/j.jhep.2024.03.031

21. Hajian-Tilaki K. Sample size estimation in diagnostic test studies of biomedical informatics. J Biomed Inform. Apr 2014;48:193–204. doi:10.1016/j.jbi.2014.02.013

22. DeLong ER, DeLong DM, Clarke-Pearson DL. Comparing the areas under two or more correlated receiver operating characteristic curves: a nonparametric approach. Biometrics. Sep 1988;44(3):837–45.

