## Supplemental Materials for "Clinical Sentiment Analysis by Large Language Models Enhances the Prediction of Hepatorenal Syndrome in Decompensated Cirrhosis"

**Affiliations:**

| **Supplemental Materials A: Participant characteristics by test/train split** | | | |
| --- | --- | --- | --- |
|  | **Test (N=59)** | **Train (N=255)** | **Overall (N=314)** |
| **Age** |  |  |  |
| Median [Q1, Q3] | 58.8 [52.5, 64.0] | 59.5 [52.3, 64.1] | 59.5 [52.3, 64.1] |
| **Race/Ethnicity** |  |  |  |
| Asian/Pacific Islander | 4 (6.8%) | 20 (7.8%) | 24 (7.6%) |
| Black/African American | 3 (5.1%) | 6 (2.4%) | 9 (2.9%) |
| Hispanic or Latino | 19 (32.2%) | 105 (41.2%) | 124 (39.5%) |
| Unknown/Other | 5 (8.5%) | 14 (5.5%) | 19 (6.1%) |
| White | 28 (47.5%) | 110 (43.1%) | 138 (43.9%) |
| **Sex at Birth** |  |  |  |
| Female | 29 (49.2%) | 122 (47.8%) | 151 (48.1%) |
| Male | 30 (50.8%) | 133 (52.2%) | 163 (51.9%) |
| **Ascites** |  |  |  |
| Count (proportion) | 20 (33.9%) | 89 (34.9%) | 109 (34.7%) |
| **Baseline Creatinine** |  |  |  |
| Median [Q1, Q3] | 1.13 [0.86, 1.43] | 1.11 [0.87, 1.63] | 1.12 [0.87, 1.59] |
| **Albumin on Admission** |  |  |  |
| Median [Q1, Q3] | 2.90 [2.30, 3.55] | 2.80 [2.30, 3.40] | 2.85 [2.30, 3.40] |
| **Total Bilirubin on Admission** |  |  |  |
| Median [Q1, Q3] | 6.0 [2.6, 12.8] | 4.6 [2.6, 10.4] | 4.65 [2.6, 10.9] |
| **MELD 3.0** |  |  |  |
| Median [Q1, Q3] | 32 27, 36] | 31 [28, 36] | 31 [28, 36] |
| **MAP on Admission** |  |  |  |
| Median [Q1, Q3] | 79 [69, 90] | 78 [70, 87] | 78 [70, 88] |
| **Sentiment Value** |  |  |  |
| Median [Q1, Q3] | 0.7 [0.6, 0.85] | 0.7 [0.6, 0.85] | 0.7 [0.6, 0.85] |
| **Refractory Ascites** |  |  |  |
| Count (proportion) | 30 (50.8%) | 132 (51.8%) | 162 (51.6%) |
| **Diuretic Intolerance** |  |  |  |
| Count (proportion) | 33 (55.9%) | 129 (50.6%) | 162 (51.6%) |
| **Shock** |  |  |  |
| Count (proportion) | 4 (6.8%) | 28 (11.0%) | 32 (10.2%) |

| **Supplemental Materials B: Performance of LLM for diagnosis of HRS compared to manual chart review** | |
| --- | --- |
|  | gpt-4o LLM |
| Sensitivity | 0.744 |
| Specificity | 0.949 |
| PPV | 0.821 |
| NPV | 0.923 |
| AUC | 0.847 |
| F1 score | 0.780 |

**Supplemental Materials C:**

System prompt: "You are an analyst who specializes in both kidney and liver disease, and in particular have expertise in identifying patients at risk of hepatorenal syndrome. You also have a background in sentiment analysis.”

**Supplemental Materials D:**

User prompt: “I will give you an unstructured clinical note taken from patients with cirrhosis who are hospitalized with an acute kidney injury on the day of their admission. I want you to read the note and give a number ranging from 0 to 1 (where 0 is no suspicion at all, and 1 is extremely suspicious) based on the sentiment analysis of the note and how strongly the clinical team suspects this patient has hepatorenal syndrome. Walk me through your reasoning for your score. I want you to give me 5 unique words from the assessment that explain your reasoning for the score. These terms should not overlap with one another. I also want you to answer the following questions with yes or no: 1. Does the patient have refractory (defined as ascites that recurs shortly after paracentesis) or resistant (ascites that is not manageable by diuretics alone) ascites? 2. Does the patient have a history of not tolerating diuretics due to recurrent kidney injury? 3. Is there concern for shock including the use of vasopressors? I want you to start your response by explaining your logic, and then end with the following structure: HRS sentiment: XX, Term1: XX, Term2: XX, Term3: XX, Term4: XX, Term5: XX, Q1: yes/no, Q2: yes/no, etc.”

**Supplemental Materials E:**

User prompt: “I am going to give you a series of unstructured discharge summaries. Each of these patients has decompensated cirrhosis and had AKI at the time of admission. Given that HRS-AKI is difficult to diagnose, we also rely on clinicians to detect and diagnose HRS-AKI, which may be commented on in the discharge summary. If the discharge summary states that the etiology or suspected etiology of the AKI is HRS-AKI, then we treat that as evidence of the etiology of AKI. I want you to tell me yes if the patient had AKI secondary to hepatorenal syndrome, or no if the patient's AKI was from an alternate etiology.”
